# Epigenetic Signatures of Human Myocardium and Brown Adipose Tissue Revealed with Simultaneous Positron Emission Tomography and Magnetic Resonance of Class I Histone Deacetylases

**DOI:** 10.1101/2020.12.06.20244814

**Authors:** David Izquierdo-Garcia, Jacob M. Hooker, Frederick A. Schroeder, Choukri Mekkaoui, Tonya M. Gilbert, Marcello Panagia, Cheryl Cero, Lindsey Rogers, Anisha Bhanot, Changning Wang, Aaron M. Cypess, Ciprian Catana, David E. Sosnovik

## Abstract

**Rationale:** Histone deacetylases (HDACs) play a central role in cardiac hypertrophy and fibrosis in preclinical models. However, their impact in the human heart remains unknown.

**Objective:** We aimed to image HDAC expression in the human heart *in vivo* with PET-MR (positron emission tomography and magnetic resonance) using [^11^C]Martinostat, a novel radiotracer targeted to class I HDACs. We further aimed to compare HDAC expression in the heart with its expression in skeletal muscle and brown/white adipose tissue (BAT/WAT).

**Methods and Results:** The specificity and selectivity of [^11^C]Martinostat binding in the heart was assessed in non-human primates (n=2) by *in vivo* blocking studies and with an *ex vivo* cellular thermal shift assay (CETSA) of HDAC paralog stabilization by Martinostat. PET-MR imaging of [^11^C]Martinostat was performed in healthy volunteers (n=6) for 60 minutes to obtain time-activity curves of probe uptake and kinetics. qPCR of class I HDACs was performed in specimens of BAT obtained from patients (n=7) undergoing abdominal surgery and in specimens of human subcutaneous WAT (n=7). CETSA and the blocking studies demonstrated that Martinostat was specific for class I HDACs in the heart. HDAC density, measured by standardized uptake values of [^11^C]Martinostat, was 8 times higher in the myocardium than skeletal muscle (4.4 ± 0.6 vs. 0.54 ± 0.29, p<0.05) and also significantly higher in BAT than WAT (0.96 ± 0.29 vs. 0.17 ± 0.08, p<0.05). qPCR confirmed higher class I HDAC expression in BAT, particularly HDAC2 and HDAC3 (2.6 and 2.7-fold higher than WAT respectively, p<0.01).

**Conclusions:** Class I HDAC expression in the human heart can be imaged *in vivo* and is dramatically higher than any other peripheral tissue, including skeletal muscle. The high levels of HDAC in the myocardium and BAT suggest that epigenetic regulation plays an important role in tissues with high energetic demands and metabolic plasticity.

## Introduction

Histone deacetylases (HDACs) are a family of enzymes that, among other functions, catalyze the hydrolysis of acetyl-L-lysine to free L-lysine side chains in protein substrates. Certain members of the family operate within an epigenetic capacity by altering chromatin remodeling and thus gene transcription in mammalian cells.^1^ Three classes of zinc-dependent HDACs exist,^2^ and a subset in class I, HDAC paralogs 1, 2, and 3, have been strongly implicated with cardiovascular disease.^3-8^ Preclinical studies have shown that the inhibition of class I HDACs can prevent, and even reverse, left ventricular hypertrophy (LVH) and myocardial fibrosis.^9-11^ However, the role of HDACs in the adult human heart remains poorly defined and difficult to assess. In addition, the expression of HDAC in other metabolically active tissues, such as skeletal muscle and brown adipose tissue, has not been well characterized.

HDAC expression in the brain and peripheral organs can be imaged *in vivo* with the positron emission tomography (PET) radiotracer, [^11^C]Martinostat.^12,13^ Here, we aimed to use this radiotracer to assess relative HDAC expression in the human heart *in vivo*, and to determine how this compares to HDAC expression in other tissues with high energetic demands. Positron emission tomography and magnetic resonance (PET-MR) imaging of [^11^C]Martinostat uptake was therefore measured in the heart, bone marrow, skeletal muscle, white adipose tissue (WAT) and brown adipose tissue (BAT) in 6 healthy volunteers. In addition, the mRNA expression of class I HDACs was assessed in specimens of BAT, acquired from 7 patients undergoing abdominal surgery. These studies, to the best of our knowledge, provide the first measurement of HDAC in human BAT and the first successful report of HDAC imaging in the human heart i*n vivo*.

## Materials and Methods

[^11^C]Martinostat dynamics and regional distribution have been determined in the young, healthy human brain, where the tool appears sensitive and specific for HDAC expression.^12-16^ The binding of [^11^C]Martinostat to class I HDACs in the non-human primate myocardium was demonstrated in this study in three steps: 1) A cellular thermal shift assay (CETSA) was performed on post mortem myocardial specimens from the hearts of non-human primates, 2) the uptake of [^11^C]Martinostat in the myocardium of a non-human primate was blocked by pre-injection of the non-radioactive isotopalogue and, 3) the uptake of ^11^C-Martinostat in the myocardium was blocked by the infusion of a competitive inhibitor of class I/IIb HDACs (suberoylanilide hydroxamic acid, SAHA). All animal studies were performed under a protocol approved by our Institutional Animal Care and Use Committee. In addition to its distribution in non-human primates, the uptake dynamics of [^11^C]Martinostat were also determined in myocardium of healthy human volunteers.

### Cellular Thermal Shift Assay

Myocardial tissue from non-human primates *(Papio anubis, n=2)* was removed from storage at - 80 ° C, 50-100 mg was cut while frozen and sealed in a custom bag (∼ 2.5 x 4 cm^2^) made from temperature stable plastic (Kapak by Ampak, 2.5 mm thickness), deeply frozen by submerging in liquid nitrogen for 5 min and manually pulverized for 20-30 sec between the flat side of an aluminum block and a 12 ounce hammer with a metal head, both pre-chilled on dry ice. Tissue was suspended by adding lysis buffer containing PBS, 0.15% NP-40, and a protease inhibitor cocktail (Roche 04693159001) at 50 mg/ml directly to the plastic bag. The tissue suspension was transferred to a 1.5 mL Eppendorf tube and homogenized with an electric pestle (2 min) then sonicated on ice at 50% power (Fisher Scientific Model CL-18) using thirty 1-second pulses with a 1 second interval between pulses. The lysate was triturated on ice with a disposable 5 mL syringe fitted with a 23 ga needle (10 passages, 2 min total) avoiding introduction of air/foam, then incubated with rotation at 4 °C for 20 min, and centrifuged at 18,000 x *g* at 4 °C for 20 min. Supernatant was collected and total protein concentration was measured using a bicinchoninic acid (BCA) protein assay (Pierce 23227).

To determine the thermal stability of HDAC paralogs in myocardial tissue, lysate was heated in three degree increments from 34 °C to 69 °C for 3 min in a thermocycler and then cooled at 10 °C for 3 min. Aliquots were immediately centrifuged (18,000 x *g* at 4 °C) for 10 min. Supernatants were then collected, 3X SDS-PAGE loading buffer containing 125 mM DTT was added, samples were heated to a boil for 10 min, and western blotting was performed. Relative mean band intensity was plotted and fitted with the Boltzmann sigmoidal equation (Prism, Graphpad) to obtain melting temperatures (Tm). Based on the Tm, 60 °C was chosen for HDAC paralogs 1/2/6 assessment and 65°C was chosen for HDAC3 assessment in the concentration gradient CETSA.

Martinostat binding to HDAC paralogs in myocardial tissue was measured by dividing the lysates into 115 μL aliquots and adding non-radiolabeled Martinostat at the following doses (0, 0.026, 0.13, 0.64, 3.0, 16, 80, 400, 2,000, and 10,000 nM). The Martinostat-containing aliquots were incubated for 30 min at room temperature with rotation, sub-divided into 50 μL aliquots, heated to 60 °C or 65 °C for 3 min, and at cooled at 10 °C for 3 min. The aliquots were then immediately centrifuged (18,000 x *g* at 4 °C) for 10 min, supernatants were collected, 3X SDS-PAGE loading buffer containing 125 mM DTT was added, samples were boiled for 10 min, and western blotting was performed. Reference (unheated) aliquots did not contain Martinostat and were kept at room temperature. Lysates were resolved by Criterion Stain-Free 4-20% gels (Biorad 567-8095) and western blotting was performed with the following antibodies: HDAC1: PA1-860 1/1000 (Thermo Fisher); HDAC2: ab124974 1/5000 (Abcam); HDAC3: ab32369 1/5000 (Abcam); and HDAC6: sc11420 1/2000 (Santa Cruz).

Thermal stabilization of HDAC paralogs conferred by incubation with a given concentration ‘x’ of Martinostat (nM) was calculated from immunoreactive band intensity (IR) as follows:

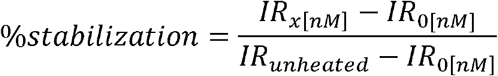

The average % stabilization (*n*= 2 biological replicates) of each HDAC paralog was calculated and plotted as an average heat map.

### Non-radioactive Martinostat Competition Study

To demonstrate that the uptake of [^11^C]Martinostat in the myocardium occurred through a specific and saturable mechanism, a competition study was performed in a non-human primate by pre-injection of the non-radioactive isotopologue, as previously reported.^13^ Martinostat (0.5 mg/kg) was co-injected intravenously with 4 mCi of [^11^C]Martinostat and PET images were acquired on a 3T MR-PET system (Biograph mMR, Siemens Healthineers, Erlangen, Germany) for 80 minutes. These data were compared to those obtained from a study without any blocking, performed on the same animal, in which 4 mCi of [^11^C]Martinostat was injected. Time activity curves (TACs) of the percent-injected dose (%ID) in the myocardium were generated for both the blocked and unblocked studies and the difference between the curves was calculated.

### Suberoylanilide hydroxamic acid (SAHA) Blocking Study

The selectivity of [^11^C]Martinostat uptake for class I HDACs in the myocardium was further demonstrated, as previously reported,^13^ by a competition study using SAHA (Vorinostat, Merck, USA), a well-established inhibitor of class I and class IIb HDACs. A baseline PET-MR scan was acquired in an adult female *Papio anubis* baboon following intravenous administration of 3.0 mCi of [^11^C]Martinostat. After 45 minutes of dynamic baseline PET scanning, a contralateral infusion of SAHA (Vorinostat) was given at a rate of 1.0 mg/kg/hr. Seventy-five minutes after the start of the SAHA infusion, a second bolus of 3.1 mCi [^11^C]Martinostat was administered followed by105 minutes of dynamic PET imaging. The infusion of SAHA was maintained for 2.5 hours. It should be noted that the preparation of SAHA (Vorinostat) used in this experiment had to be dissolved in a pH 12 solution for IV injection, limiting the dose that could be used in the blocking study.

### Human Imaging Studies

Six healthy volunteers (3 females and 3 males, mean age ± SD: 42.7 ± 15.5 years) participated in the human imaging pilot study. The participants had no major medical conditions and, in particular, no history of diabetes, hypertension or cardiovascular disease. All subjects provided written consent to participate in the study under a protocol approved by the Institutional Review Board at the Massachusetts General Hospital and occurring under FDA eIND #123154.

### Image Acquisition

[^11^C]Martinostat was synthesized as previously described.^16^. Simultaneous PET and MR data were acquired on the Biograph mMR scanner (Siemens, Erlangen, Germany). The probe was injected intravenously with the patients in the scanner, and PET data were acquired in 3D list-mode for 60 min following injection. ECG and respiratory waveforms were recorded throughout the acquisition and incorporated into the list-mode data. Approximately 150 MBq (mean ± SD: 151.1 ± 33.9 MBq) of [^11^C]Martinostat was administered. No physiological changes or adverse reactions were noted with [^11^C]Martinostat administration.

Cardiac MR images were acquired simultaneously during the PET acquisition using a flexible 6-channel coil array. Balanced steady state free precession (bSSFP) cines were performed in the short axis of the entire left ventricle (LV) with the following parameters: Field of view (FOV)=292×360 mm^2^, matrix size= 256×208, slice thickness=7 mm with no slice gap, repetition time/echo time (TR/TE)=3/1.5 ms, flip angle=48°, readout bandwidth=977 Hz/pixel, retrospective ECG gating, 16 lines of k-space per acquisition window, number of reconstructed cardiac phases=25.

Attenuation correction of the PET data was performed using a dual-echo three dimensional Dixon-VIBE (Volume Interpolated Breath-hold Examination) sequence, acquired in the coronal plane, with the following parameters: FOV=500×328×396 mm^3^, matrix size=192×126×128, voxel size=2.6×2.6×3.12 mm^3^, TR=3.6 ms, TE=1.23/2.46 ms, flip angle=10°, acquisition time=18 sec. Dixon images were acquired during breath-hold at end expiration to match the reconstructed PET images (see below) and therefore minimize the attenuation-emission mismatch due to respiratory motion.

### Image Processing

Cardiac mass, volume and ejection fraction were calculated from the cine images on a Syngo workstation (Siemens, Erlangen, Germany). The dynamic PET images were reconstructed from the list-mode data using the following framing: 12×5sec, 6×10sec, 6×30sec, 5×1min, 5×2min and 8×5min. Images were reconstructed using a 3D Ordinary-Poisson Ordered Subset Expectation-Maximization (OP-OSEM) algorithm, with 3 iterations and 21 subsets, and with corrections applied for attenuation, isotope decay, dead time, normalization, sensitivity, scatter and random coincidences. [^11^C]Martinostat kinetics were assessed in regions of interest (ROIs) over the myocardium, blood pool (inside the LV), liver, kidneys and lungs. TACs were obtained in each ROI by calculating the mean and standard deviation of the standardized uptake values (SUVs),^17^ across subjects on each individual dynamic PET frame and plotting them over time. The ratio of probe uptake in the myocardium/blood (RM/B) was also calculated and plotted over time in a similar fashion.

Static PET images were also reconstructed from the data acquired from 10 to 30 minutes after injection, using retrospective gating at end expiration. Mean SUVs were calculated from these static PET images in the myocardium, skeletal muscle, bone marrow (thoracic vertebrae), subcutaneous WAT and paravertebral BAT. Vertebrae in close proximity to the liver were excluded to avoid partial volume effects. Foci of paravertebral BAT were identified on the Dixon fat images using a two-step approach as previously described.^18^ In each voxel the water/fat ratio was calculated and those voxels with a ratio <0.15 were defined as WAT, those > 0.85 as bone marrow and those in the paravertebral fat with a ratio of 0.15-0.85 were designated as BAT. Care was taken to ensure that no portion of the BAT ROI overlapped with the aorta or ribs, which were easily identified on the axial Dixon images. Fusion of the PET and MR data was performed offline using the OsiriX DICOM viewer (University of Geneva). Statistical comparison of SUVs (mean and standard deviation) was performed with a Mann-Whitney test in the following tissues: myocardium versus skeletal muscle and BAT versus WAT.

### HDAC Expression in Human Adipose Tissue

The mRNA expression of class I HDACs in human BAT and WAT was measured with quantitative real-time PCR (qPCR). Samples of human paravertebral and peri-adrenal BAT were harvested from patients (n=7) undergoing abdominal (adrenal) surgery and compared to samples of human subcutaneous WAT obtained during autopsy (n=7). Total RNA was extracted from the tissue samples using an RNeasy minikit (Qiagen). First strand cDNA was synthesized from 1ug of total RNA using the High Capacity cDNA Reverse Transcription kit (Applied Biosystems). Quantitative RT-PCR was performed on the ABI Prism 7900 Sequence Detection System (Applied Biosystems, Carlsbad, Calif.) using Taqman assays Hs00606262_g1 for HDAC1, Hs00231032_m1 for HDAC2, Hs00187320_m1 for HDAC3, Hs0022453 for UCP1 (Applied Biosystems) Relative mRNA expression was determined by the 2^—ΔΔCt^ analysis method using β actin (Hs_99999903_m1) as an endogenous control. UCP1 and HDAC levels were normalized to the values in WAT and compared with a Mann-Whitney test. Fold-differences (mean ± standard deviation) between BAT and WAT are reported.

## Results

### Binding of Martinostat to Class I HDACs

CETSA revealed that Martinostat stabilized HDAC1, HDAC2 and HDAC3 in the myocardium, but not HDAC6 (Figure 1). Thermal stabilization for the HDAC paralogs 1, 2, and 3 was observed with increasing concentrations of Martinostat. For comparison, 35% stabilization was achieved at a Martinostat concentration of 400 nM for HDAC1, 2,000 nM for HDAC2, and 10,000 nM for HDAC3. These results demonstrate that Martinostat is capable of engaging class I HDAC paralogs (1, 2, and 3) in a preparation of myocardial tissue, and supports the utility of [^11^C]Martinostat to image class I HDACs in the heart with PET-MR.

**Figure 1:**
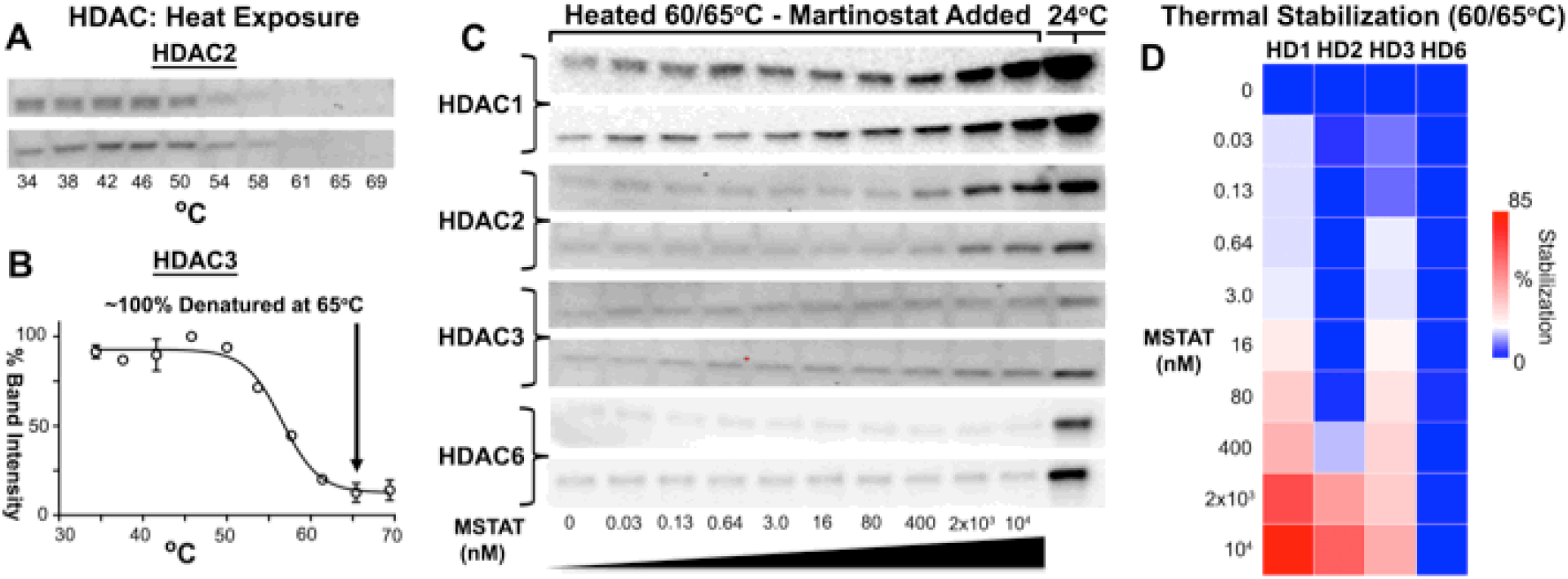
Cellular thermal shift assay (CETSA) of Martinostat HDAC engagement in the myocardium. Whole cell lysates were prepared from *Papio anubis* myocardium (*n*=2 baboons). The melting temperatures (T_m_) of HDAC paralogs were determined through western blotting and relative band intensities were fitted to a sigmoidal Boltzmann curve. (A) Representative western blot images from HDAC2, which has a T_m_ of 54 °C. (B). Representative melting curve from HDAC 3, which has a T_m_ of 57 °C. Based on the T_m_ results, 60° C was chosen to assess stabilization of HDAC paralogs 1, 2, and 6, while 65°C was chosen to assess stabilization of HDAC3. (C) The thermal stabilization of HDAC paralogs 1, 2, 3, and 6, in the presence of increasing concentrations of Martinostat, was compared through western blotting. Martinostat thermally stabilizes class I HDACs (paralogs 1, 2 and 3). (D) Heat map of average % thermal stabilization shows that Martinostat stabilizes class I HDACs but has no detectable effect on HDAC6 (class IIb) in this assay. MSTAT = Martinostat; HD = HDAC

The results of the competition imaging studies are shown in the supplement. Re-analysis of the blocking data in the current study was performed by reformatting the images into the short axis plane of the heart, ensuring that the regions-of-interest used were placed in an optimal location in the myocardium. The co-injection of non-radioactive Martinostat reduced the percent-injected dose (%ID) in the myocardium by 75%. Infusion of SAHA reduced [^11^C]Martinostat uptake in the myocardium. The percent injected dose of [^11^C]Martinostat blocked by SAHA reached 50% in the heart, which reflects the specificity of [^11^C]Martinostat for cardiac HDACs and the limited dose of SAHA that could be safely injected. The uptake of the probe in the liver, in contrast, was non-specific and increased with SAHA infusion. This likely reflects the increased availability of [^11^C]Martinostat to the liver due to its uptake in other organs being blocked by SAHA.

### Uptake of ^11^C-Martinostat in Human Myocardium

Robust uptake of [^11^C]Martinostat in the LV was consistently seen in all 6 healthy volunteers (Figure 2). Low levels of background uptake were seen in the blood pool and lungs, making myocardial uptake of the probe very conspicuous. TACs showed that SUVs in the blood pool dropped very rapidly in the first 10 minutes after injection, and then decreased very slowly (Figure 2). In contrast, SUVs in the myocardium exhibited a monoexponential decay pattern, and averaged 4.4 ± 0.6 10-30 minutes after probe injection. The ratio of SUVs in the myocardium/blood averaged 1.9 ± 0.4 during the 10-30 minute time window and remained relatively stable until the end of the scan (Figure 2).

**Figure 2:**
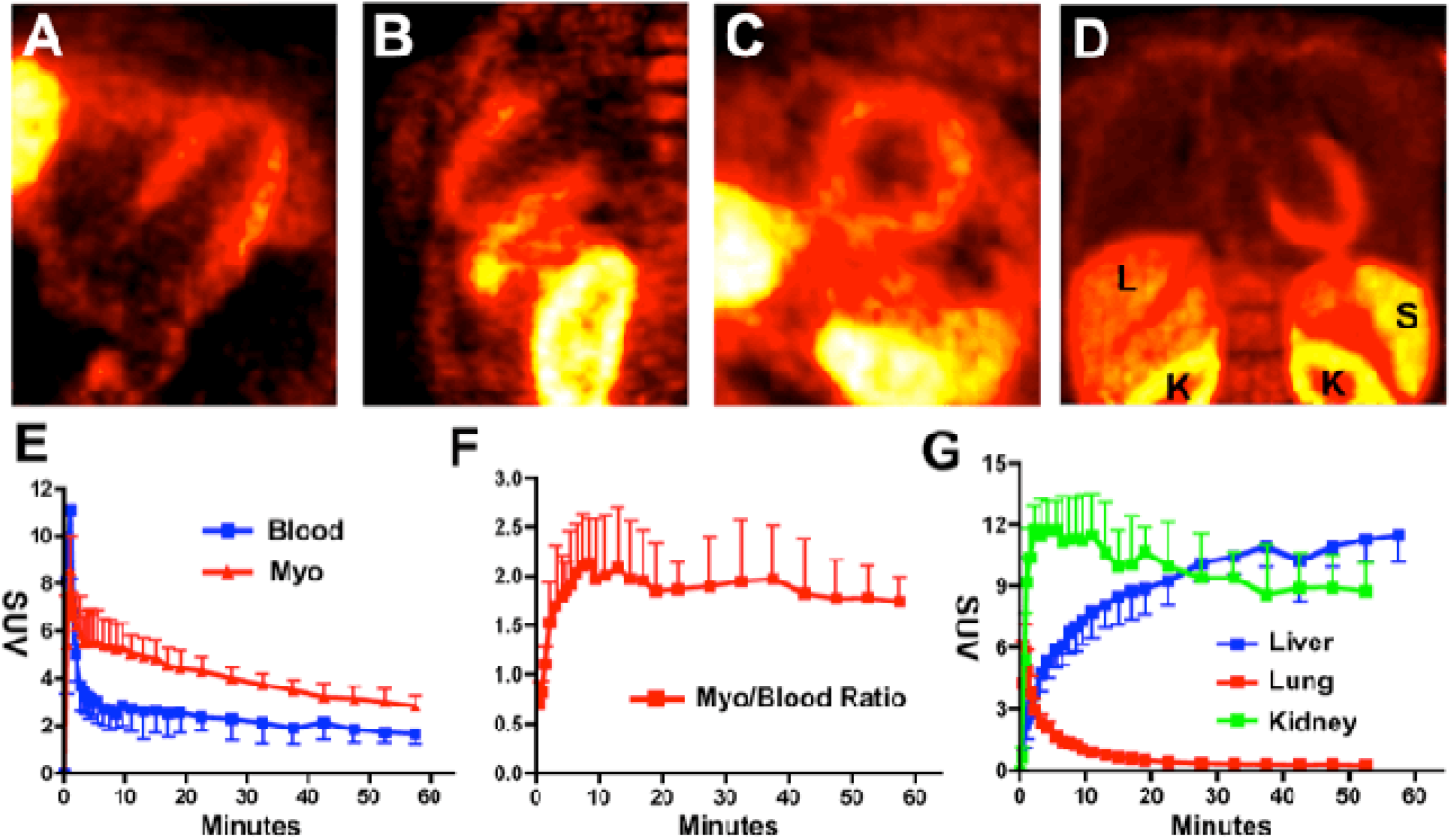
Time activity curves of [^11^C]Martinostat in the human heart and vital organs. Multiplanar reformats 10-30 minutes after injection are shown in the 4-chamber (A), 2-chamber (B), and short-axis (C) views of the left ventricle and in the coronal plane (D). Significant accumulation of the probe occurs in the kidneys (K) and spleen (S) within 30 minutes. However, the signal in the liver (L) is low. (E) Time activity curves from the myocardium and blood 0-60 minutes post [^11^C]Martinostat injection. Washout of the probe from the blood pool is very rapid and near-complete within 10 minutes. In contrast, probe accumulation in the myocardium is significant and remains elevated even 60 minutes after probe injection. (F) The ratio between [^11^C]Martinostat uptake in the myocardium and blood reaches approximately 2 within ten minutes of probe injection and remains elevated for 60 minutes. (G) Time activity curves from the liver, lung, and kidney 0-60 minutes post [^11^C]Martinostat injection. The rapid washout of the probe in the lungs and slow accumulation in the liver creates a high-target to background ratio in the heart 10-30 minutes after injection.

Integration of the PET and cine MR datasets was successfully performed in all six volunteers (Figure 3). No correlations were seen between [^11^C]Martinostat uptake and any of the standard physiological parameters in the cine images (see supplement), which all fell within the normal range. Our data suggest that HDAC expression levels in the heart may be higher in females but a further study with a larger number subjects will be needed to assess this. Volume rendered images of [^11^C]Martinostat uptake in the LV are shown in Figure 4. The cine MR data were used to create a 3D template of the heart, which was spatially co-registered with the PET data. The distribution of [^11^C]Martinostat in all myocardial segments was similar, however, a gradient in signal from the endocardium (highest) to epicardium (lowest) was frequently seen.

**Figure 3.**
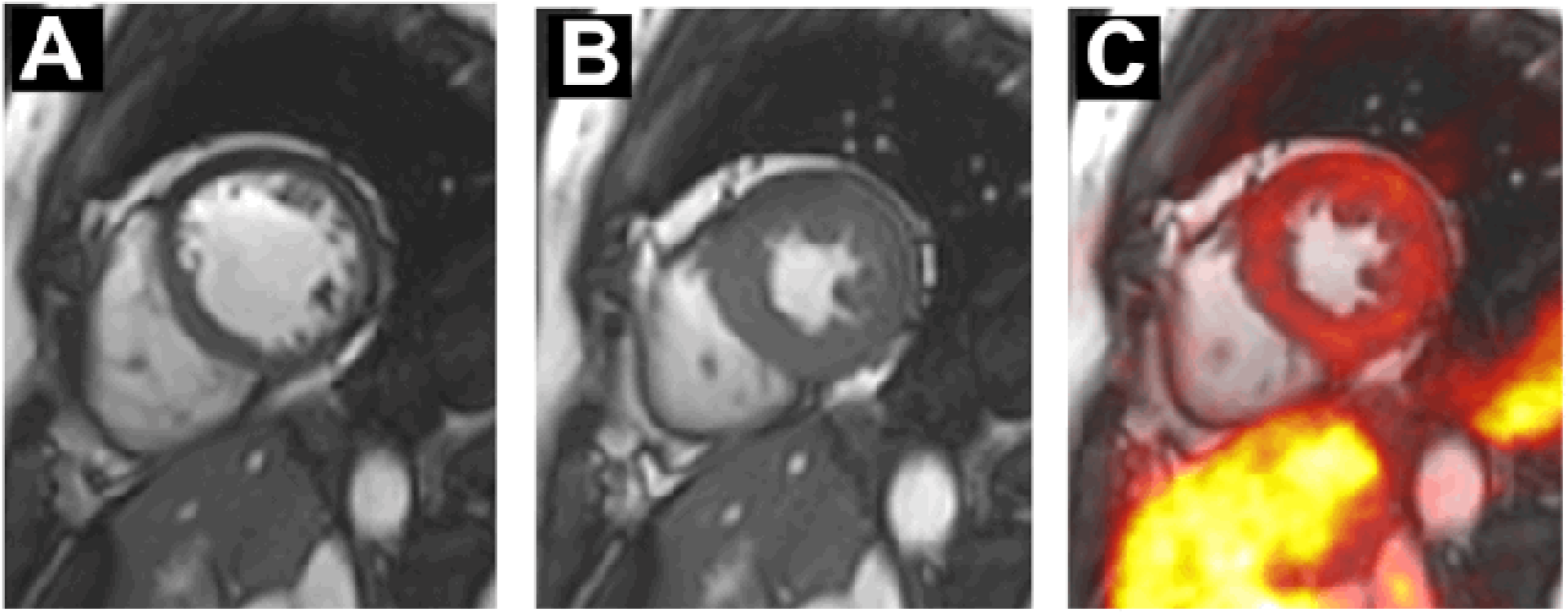
Simultaneous cardiac PET-MR imaging of HDAC expression in the human heart. MR cine frames from (A) end diastole and (B) end systole are shown. (C) Fusion of a short axis PET image and an end-systolic MR image. The uptake of [^11^C]Martinostat in the myocardium and liver can be well visualized.

**Figure 4.**
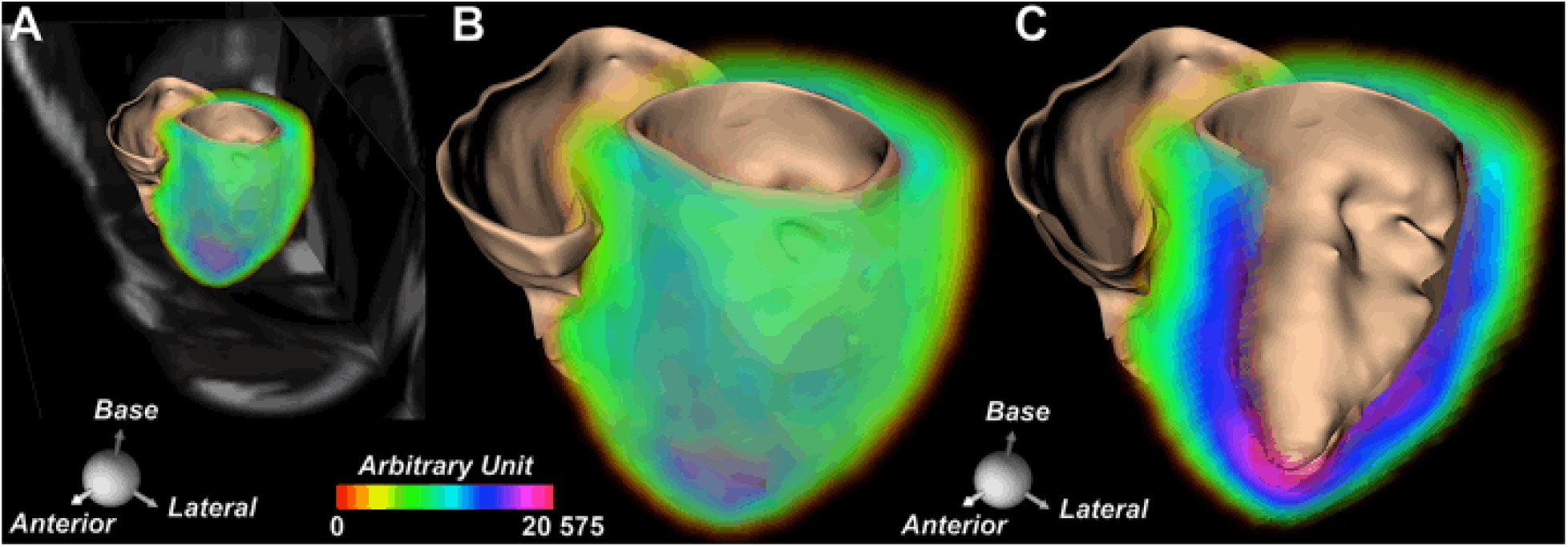
Volume rendered images of [^11^C]Martinostat uptake in the human heart. (A, B) The cine MR images have been used to form an anatomical template of the heart and co-registered with the PET images of probe uptake in the myocardium. The distribution of the probe in most myocardial segments is extremely similar. However, removal of the anterior wall (panel C) reveals a transmural gradient in the PET signal from endo to epicardium, which is likely related to cardiac motion and underscores the potential of PET-MR to further enhance PET images through advanced motion correction strategies.

### HDAC Expression in Extracardiac Tissues

Foci of high [^11^C]Martinostat signal were consistently seen in areas of paravertebral BAT (Figure 5). Use of the fat, water and ratio Dixon images allowed ROIs to be drawn in BAT, while avoiding volume averaging with adjacent tissues. A high degree of co-localization was seen between areas of paravertebral BAT on Dixon MRI and paravertebral foci of [^11^C]Martinostat uptake (Figure 5). [^11^C]Martinostat uptake in the other extra-cardiac tissues varied significantly, ranging from lowest in WAT to highest in the bone marrow. Specifically, the SUV of [^11^C]Martinostat in the bone marrow was (1.55 ± 0.32), which was markedly lower than the SUV in the myocardium (Figure 6). Analysis of TACs in the bone marrow of the non-human primates (Figure 5) confirmed that the uptake of the probe in the marrow was specific.

**Figure 5.**
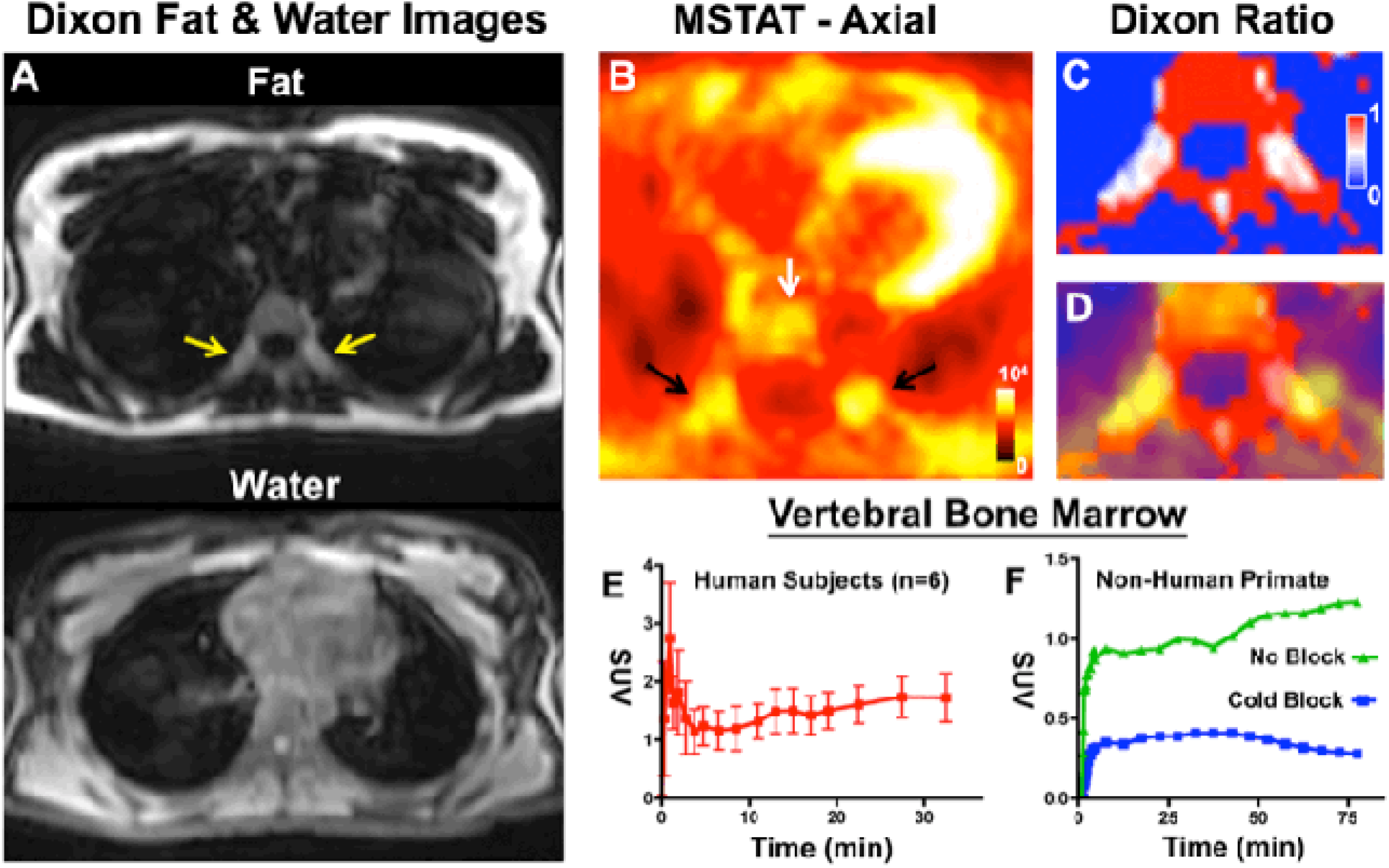
[^11^C]Martinostat uptake in human extracardiac tissues. (A) Dixon fat and water images showing foci of paravertebral fat (arrows) that also have intermediate intensity on the water image. (B) [^11^C]Martinostat (MSTAT) uptake in the vertebral bone marrow (white arrow) and paravertebral fat (black arrows) is high. (C) Dixon ratio image: Foci of BAT are characterized by intermediate water/fat ratios (>0.15 and < 0.85; pale blue to pale red). (D) Fusion of Dixon ratio and [^11^C]Martinostat images confirms the high uptake of the probe in paravertebral BAT. (E) Time activity curve of [^11^C]Martinostat uptake in the bone marrow in 6 healthy volunteers 0-30 minutes post [^11^C]Martinostat injection. (F) Time activity curves of bone marrow in non-human primate with baseline and preinjection of cold compound 0-80 minutes post [^11^C]Martinostat injection. Pre-injection of cold Martinostat in a non-human primate blocks probe uptake in the bone marrow, demonstrating that uptake is specific in this tissue.

**Figure 6.**
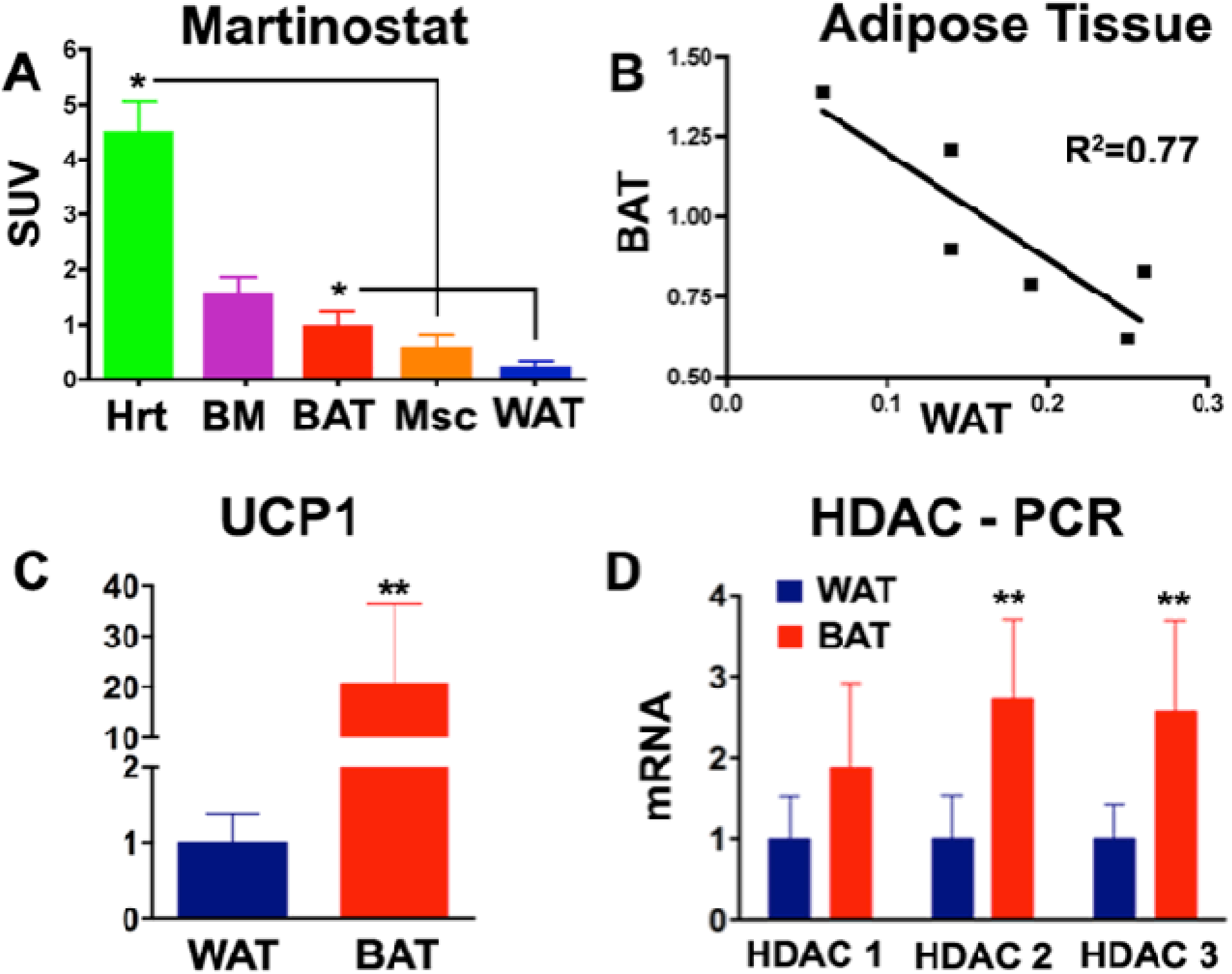
Class I HDAC expression in human BAT. (A) The uptake of [^11^C]Martinostat was significantly higher in BAT than WAT (p<0.05), and probe uptake in the heart was 8-fold higher than in skeletal muscle (p<0.05). (B) An inverse correlation was seen between [^11^C]Martinostat uptake (SUVs) in BAT and WAT. (C) qPCR was used to detect mRNA levels of *UCP-1* and class I HDACs *(HDAC1, HDAC2*, and *HDAC3)* across tissues, and expression was normalized by *ACTN*. Biopsy specimens of human BAT and WAT (n=7 each) show high levels of *UCP-1* expression in BAT compared to WAT, confirming tissue identity. Expression of *HDAC2* and *HDAC3* was significantly higher in BAT compared to WAT. (Hrt = heart, BM = bone marrow, Msc = skeletal muscle). * p<0.001; ** p<0.01.

Striking differences in probe uptake were also seen between tissues belonging to the same overall tissue class. For instance, significant differences in [^11^C]Martinostat SUVs were seen between the myocardium and skeletal muscle (4.4 ± 0.6 vs. 0.54 ± 0.29, p<0.05), and between BAT and WAT (0.96 ± 0.29 vs. 0.17 ± 0.08, p<0.05). In addition, an inverse relationship (R^2^ = 0.77, p<0.05) was seen between [^11^C]Martinostat uptake in BAT and WAT (Figure 6).

qPCR of samples of human adipose tissue showed that the mRNA expression of class I HDACs was significantly higher in BAT than WAT (Figure 6), supporting the *in vivo* imaging results. Gene expression in BAT versus WAT was not significant for HDAC1 (1.9 ± 1.0 fold higher, p = 0.2), was 2.7 ± 1.0 fold higher for *HDAC2* (p=0.0023), and 2.6 ± 1.1 fold higher for *HDAC3* (p= 0.0041). Levels of UCP1 in the samples of BAT were 20.7 ± 15.6 fold higher than in WAT (p=0.0012), verifying their phenotype (Figure 6).

## Discussion

HDAC activity in preclinical models has been shown to play a major role in the development of cardiac hypertrophy and fibrosis.^9-11^ The role of HDAC in the human heart, however, has been challenging to characterize. Here, using PET-MR of a novel radiotracer we show that relative HDAC levels in human myocardium are 8-fold higher than in skeletal muscle. In addition, using PET-MR and qPCR of biopsies from human subjects, we show that relative HDAC expression is significantly higher in human BAT than WAT. Our study reveals that class I HDACs in humans are most abundantly expressed in tissues with a high degree of metabolic and microstructural plasticity.

The role of HDAC in the myocardium has been extensively explored in murine models. The inhibition of class I HDACs exerts a marked anti-fibrotic effect in the murine heart by blocking the synthesis of collagen by extracellular signal regulated kinase (ERK) and transforming growth factor (TGF).^7, 9, 10, 19-22^ HDAC inhibition can also modulate the differentiation of fibroblasts in the heart,^23^ which attenuates and even reverses myocardial fibrosis in mice.^21^ The level of HDAC expression detected by [^11^C]Martinostat in human myocardium was markedly high relative to other peripheral tissues, suggesting that similar effects might be possible in the human heart. The imaging of myocardial HDAC expression in humans could thus facilitate the exploration of therapeutic HDAC inhibition in heart failure and other cardiovascular diseases.

Skeletal muscle, unlike the myocardium, undergoes atrophy with aging, disuse and disease. Class I HDACs, and in particular HDAC1, activate the Forkhead box O (FoxO) transcription factors in skeletal muscle and regulate skeletal muscle atrophy.^24^ Interestingly, [^11^C]Martinostat uptake in skeletal muscle was far lower than in the heart, which undergoes hypertrophy rather than atrophy with age. This suggests that a significantly greater degree of epigenetic regulation may be needed to mediate a hypertrophic than an atrophic response. Recent data in rodents have shown that HDACs can also have a direct effect on cardiac myofibrils and impede their relaxation in diastole.^25^ This further underscores the potential value of [^11^C]Martinostat as readout in those with LVH and heart failure with preserved ejection fraction.

Significant differences in relative HDAC expression were also seen in this study between BAT and WAT, both by PET-MR and qPCR. In addition, in the six healthy volunteers imaged, an inverse correlation was seen between [^11^C]Martinostat uptake in BAT and WAT. This suggests that individuals with more dynamic metabolism in BAT may have more static physiology in WAT, however, further validation is required in a larger cohort.

The uptake of [^11^C]Martinostat in the bone marrow was high, consistent with the recently described role of class I HDACs in the regulation of hematopoiesis,^26^ and the production of thrombocytopenia, anemia and neutropenia by HDAC inhibitors in clinical trials.^27^ Preclinical data also suggest that class I HDACs can modulate monocyte phenotype and polarization,^28^ and the inhibition of HDAC3 has been shown to stabilize atherosclerotic plaques.^29^ The inhibition of HDAC activity in the bone marrow and blood could thus have both beneficial and deleterious effects, underscoring the need to quantify and closely monitor this process *in vivo*. Conceptually, imaging HDAC expression with [^11^C]Martinostat could allow the degree of HDAC inhibition to be rationally titrated and the beneficial effects to be maximized.

PET-MR imaging of [^11^C]Martinostat uptake in the heart provides several advantages including the ability to exploit the full range of MR contrast mechanisms to phenotype the myocardium.^30-32^ Furthermore, the use of Dixon water/fat separation techniques allows BAT to be identified based on endogenous MR contrast rather than the use of [^18^F]FDG. The PET portion of the instrument can thus be reserved for the imaging of a specific molecular tracer, such as [^11^C]Martinostat, providing a distinct advantage over PET-CT based imaging of BAT. Motion compensation of the PET data was performed in this study by accepting only those images acquired at end expiration, however, more sophisticated compensation schemes are feasible.^33^ Likewise, more sophisticated attenuation correction schemes than the one used here have recently been developed.^34^ Further work will be required to determine whether the transmural gradient in [^11^C]Martinostat uptake in the myocardium reflects the biological distribution of HDACs or whether this is an artifact produced by partial volume effects, cardiac motion, or variations in photon attenuation over the cardiac cycle.

Whole body knockout of class I HDACs is frequently lethal and, consequently, the specificity of [^11^C]Martinostat cannot be easily tested in transgenic mice. Extensive study of [^11^C]Martinostat in the brain, however, has shown that it engages class I HDACs.^12-16^ The CETSA, together with SAHA and cold blocking studies described here suggest that [^11^C]Martinostat engages class I HDACs in the myocardium as well. It should be noted, however, that a positive control for HDAC6 was not available in the CETSA experiments. Potential binding of Martinostat to HDAC6 can, therefore, not be definitively excluded. It should also be noted that the formulation of SAHA (Vorinostat) used in the blocking study needed to be dissolved in a pH 12 solution for IV injection. This limited the dose that could be safely injected *in vivo* and likely accounts for the degree of blocking with the cold-compound being higher than with SAHA (75% vs. 50%).

The key limitation of [^11^C]Martinostat lies in its lack of specificity for a single HDAC paralog. This is less of an issue in the heart where the effects of the class I HDACs, particularly HDAC1 and HDAC2, on the myocardium are similar.^6, 8^ In BAT, however, HDAC1 and HDAC3 may play divergent roles. In mice HDAC1 inhibits the thermogenic program in brown adipocytes,^35^ while HDAC3 plays a key role in the uncoupling of oxidative phosphorylation, and hence thermogenesis, in BAT.^36^ Further study of the role of HDAC in human BAT and the affinity of [^11^C]Martinostat for the relevant paralogs will be needed. Nevertheless, to the best of our knowledge, this is the first demonstration of HDAC expression levels in human BAT, either by imaging or qPCR.

## Conclusion

In conclusion, we show that HDAC expression can be imaged *in-vivo* in the human heart and in other metabolically active and plastic tissues, such as BAT. The uptake of [^11^C]Martinostat in the myocardium is markedly higher than in any other peripheral organ, underscoring the potential importance of class I HDACs in the heart. [^11^C]Martinostat produced a high target-to-background ratio in the myocardium and washout of the probe from the blood pool and lungs was rapid. The kinetics of the probe are thus well suited to cardiac imaging and clinical studies of HDAC expression in the heart could provide important insights into the role of epigenetic regulation in cardiovascular disease.

## Data Availability

All data associated with this study are available in the main text. All data can be
obtained from the authors under reasonable request.

## Acknowledgements

We thank Judit Sore and the radiopharmacy team for technical assistance in radiotracer synthesis and Grae Arabasz, Shirley Hsu, Regan Butterfield, and Marlene Wentworth for assistance with PET-MR imaging. This research was supported in part by the following National Institutes of Health grants: R01HL112831, R01HL141563 (D.E.S.), R01EB014894 (CT), R01DA030321 (J.M.H.) and P41RR14075 to the Martinos Center for Biomedical Imaging.

