## Supplemental Material for "Epigenetic Signatures of Human Myocardium and Brown Adipose Tissue Revealed with Simultaneous Positron Emission Tomography and Magnetic Resonance of Class I Histone Deacetylases"

**Supplemental Figures:**

Blocking studies in non-human primates (n=2) were performed as previously described.[1] In the current study, the images were reformatted into standard cardiac planes to facilitate optimal placement of the regions-of-interest used to calculate the standardized uptake values (SUVs).

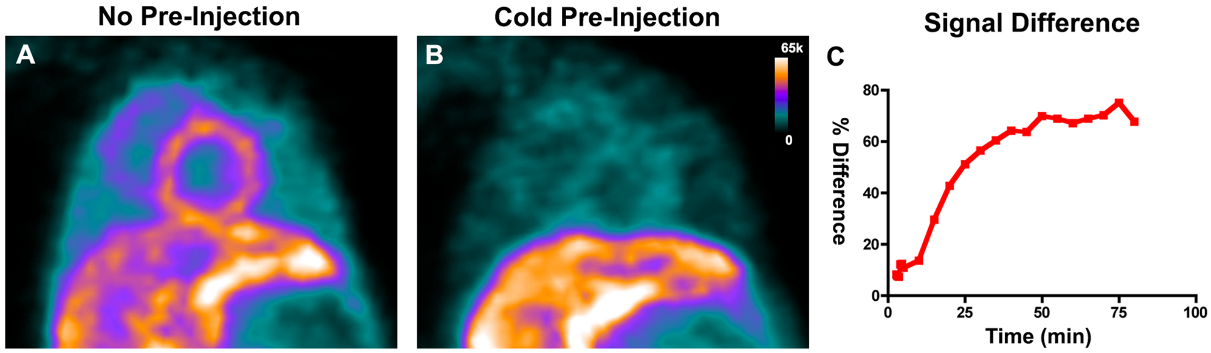


**Figure S1: Blocking of [$^{11}$C]Martinostat uptake in the heart by pre-injection of cold compound.** (A, B) Short axis images of the heart acquired (A) without blocking and (B) with blocking through pre-injection of non-radioactive Martinostat. The signal from the probe in the liver is similar. However, pre-injection of cold compound markedly attenuates the intensity of [$^{11}$C]Martinostat in the myocardium. (C) The percent-difference in the SUVs, normalized to the unblocked values, increases with time and reaches a peak of 75%.

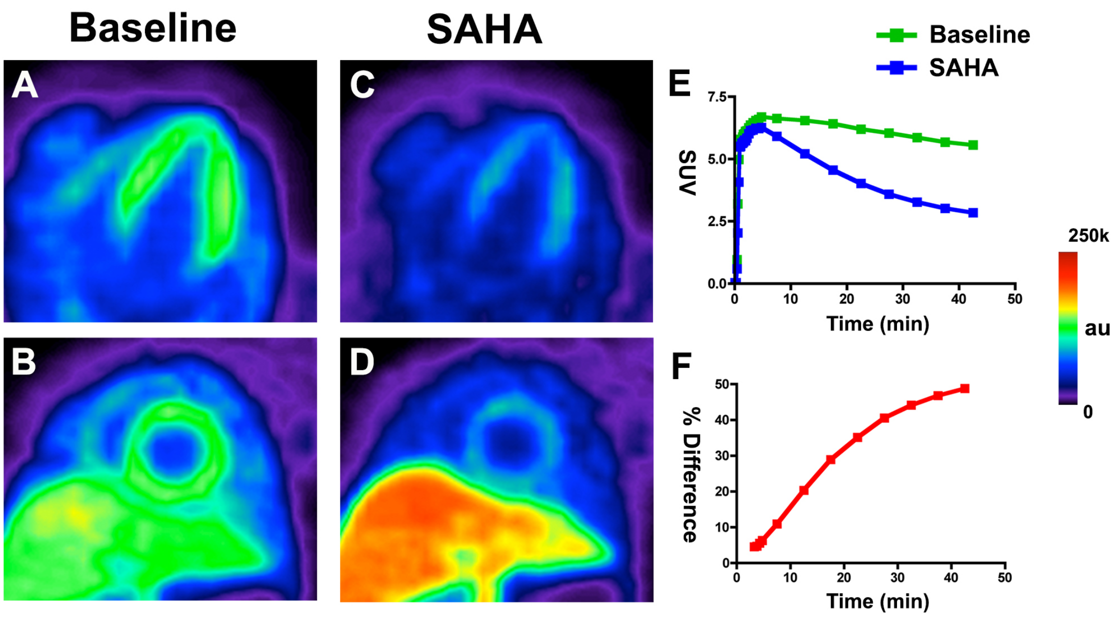


**Figure S2: Specificity of [$^{11}$C]Martinostat for myocardial HDAC demonstrated by blocking with a competitive HDAC inhibitor.** Baseline images of the heart in (A) the 4-chamber view and (B) the short axis view are shown. (C, D) Corresponding images acquired after the infusion of the competitive HDAC inhibitor suberanilohydroxamic acid (SAHA) show a marked reduction in myocardial [$^{11}$C]Martinostat signal. (E) Time activity curves from the myocardium corresponding to the baseline and SAHA-blocked images. (F) The percent-difference in the myocardial SUVs produced by pre-injection increases with time and reaches a peak of 50%. The displacement of [$^{11}$C]Martinostat produced by SAHA reflects the specificity of [$^{11}$C]Martinostat for cardiac HDACs, but is limited by the dose of SAHA that can be safely given *in vivo*. Uptake of the probe in the liver is non-specific and is not blocked by SAHA.
